# Identifying gaps in COVID-19 health equity data reporting in Canada using a scorecard approach

**DOI:** 10.1101/2020.09.23.20200147

**Authors:** Alexandra Blair, Kahiye Warsame, Harsh Naik, Walter Byrne, Abtin Parnia, Arjumand Siddiqi

## Abstract

**Objective:** To assess thealth equity-oriented COVID-19 data reporting across Canadian provinces and territories, using a scorecard approach.

**Method:** A scan was performed of provincial and territorial reporting of five data elements (cumulative totals of tests, cases, hospitalizations, deaths and population size) across three units of aggregation (province or territory-level, health regions, and local areas) (15 “overall” indicators), and for two vulnerable settings (long term care and detention facilities) and six social markers (age, sex, immigration status, race/ethnicity, essential worker status, and income) (120 “equity-related” indicators). Per indicator, one point was awarded if case-delimited data were released, 0.7 points if only summary statistics were reported, and 0 if neither was provided. Results were presented using a scorecard approach.

**Results:** Overall, information on cases and deaths was more complete than for tests, hospitalizations and population size denominators needed for rate estimation. Information provided on jurisdictions and their regions, overall, tended to be more available (average score of 53%, “B”) than for equity-related indicators (average score of 21%, “D”). Only British Columbia and Alberta provided case-delimited data, and only Alberta provided information for local areas. No jurisdiction reported on outcomes according to patients’ individual-level immigration status, race, or income. Only Ontario and Quebec provided detailed information for long-term care settings and detention facilities.

**Conclusion:** Socially stratified reporting for COVID-19 outcomes is sparse in Canada. However, several “best practices” in health equity-oriented reporting were observed and set a relevant precedent for all jurisdictions to follow for this pandemic and future ones.

## 1. Introduction

Early reporting by regional and provincial jurisdictions in Canada suggests that like in other countries such as the United States of America (USA),^1,2^ social inequities in COVID-19 outcomes have emerged in Canada. In Ontario, for instance, higher rates of COVID-19 incidence, hospitalization, and death have been observed in lower-income areas and areas with higher densities of immigrant and racialized residents.^3^ Toronto has reported that 83% of COVID-19 cases with available race data, identified between mid-May and mid-July 2020, occurred among racialized residents, despite these residents representing 52% of the city’s population.^4^

These early reports of social inequities in COVID-19 outcomes beg several questions for public health policy and intervention. Since the identification of these inequities is predicated on the availability and release of COVID-19 surveillance data according to social markers, one fundamental question is how Canada is doing, overall, in health equity-informed COVID-19 data reporting across jurisdictions? Knowing which inequities have emerged, and where, is a necessary first step in planning health equity-informed health policy and interventions.^2^

Indeed, public release and reporting on surveillance data have been essential to inform epidemiologic research and modelling and public health interventions since the start of the COVID-19 pandemic. Presenting data disaggregated by social markers, such as sex or race, can ensure that social and political responses to the health crisis are sensitive to and designed to be effective against social disparities in outcomes.^5^ Data transparency also serves to protect the public’s trust in public health guidelines and ensure accountability.^6^ However, given that provincial and territorial rather than federal public health authorities are the primary entities collecting and reporting on health data in Canada, public-facing output on local- or social marker-disaggregated data can vary across Canadian jurisdictions. An assessment of both overall and health equity-oriented COVID-19 data reporting in each Canadian province and territory is needed to identify both best practices and reporting gaps.

The objective of this study was to perform an environmental scan of COVID-19 data reporting across Canadian provinces and territories and to assess health equity-focused reporting using a scorecard approach. Scorecards can be used to help track health-related trends or the quality of data reporting across jurisdictions ^7^. Here, we build on the USA-based Coronavirus in Kids (COVKID) Tracking and Education Project’s recently proposed COVKID State Data Quality Report Card ^8^ which was designed to identify gaps in COVID-19 related surveillance in children. We propose the Canadian COVID-19 Health Equity Data Scorecard as an evaluation framework for the Canadian context.

## 2. Method

### 2.1 Data

A detailed environmental scan of official provincial and territorial public health websites and published reports was performed to identify reporting data content. Reference websites used were those provided by the Public Health Agency of Canada on their centralized reporting website.^9^ Provincial and territorial websites were searched for data summaries, figures, tables as well as downloadable reports (most often available in portable document (PDF) format), by navigating through websites and downloading and reviewing reports. The scan was performed between June 11 and August 11, 2020, and the results are accurate to the latter date.

### 2.2 Scorecard indicators

Based on the minimum data requirements proposed by extant COVID-19 data quality assessments, such as the COVKID Project Data Quality Report Card,^8^ we assessed provinces’ and territories’ reporting of five data elements: cumulative totals of tests performed, case counts, hospitalizations and deaths, as well as the availability of data on the size of populations of interest (i.e. the necessary denominator for rate estimation).

We assessed the availability of these five data elements across three geographic units of population aggregation (overall province or territory-level, health region- or unit-level, and Forward Sortation Area-level or small neighbourhood area equivalent). Reporting on various levels of spatial aggregation was assessed given that transmission epidemiology and distributions of risk factors can vary across jurisdictions. We also assessed data reporting across eight equity-related indicator strata: two settings that have been particularly vulnerable to COVID-19 outbreaks (long term care and detention facilities)^10,11^ and six social markers (age, sex, immigration status, race/ethnicity, essential worker status, and income groups). The latter social markers have been identified as key social determinants of health and infectious disease burden. ^12,13^ With the five data elements across three units of population aggregation—overall and across eight social strata, 135 indicators were used (5 * 3 * (1 “overall” stratum + 8 social strata) C= 135).

As done for other scorecards, these indicators were selected for being measurable, relevant for health equity surveillance, actionable, and interpretable.^7^ Indeed, precedent exists for surveillance reporting across all social markers used, including by race/ethnicity^14,15^ and essential worker status^16^—if not for COVID-19 than other common health outcomes.^9,17,18^

### 2.3 Analysis

For each of the 135 indicators, 1 point was awarded if raw, anonymized case-delimited data were released and publicly available (i.e. where each case represented one data row, available in a downloadable, and editable file format, such as in Comma Separated Values (.csv) format). A total of 0.7 points were awarded if summary statistics were reported for the indicator, but no raw case-delimited data were publicly available, and a score of 0 was awarded if neither information was reported nor data made publicly available. To contrast surveillance reporting across jurisdictions at a national scale, points were only awarded if the data element was available for the entire jurisdiction (i.e. not if data were only available for certain regions).

We used a near-complete (0.7 points; intentionally higher than a half-point) and complete (1 point) scoring system rather than a binary (present/absent, 0 vs. 1 point) method to acknowledge the relevance of summary statistic reporting, while rewarding jurisdictions that opted for full data transparency for public use—as done in peer nations such as the United Kingdom^19^ and the United States.^20^ Raw data sharing has been identified as a best practice in supporting innovation and research, advancing government accountability and evidence-informed decision-making. ^6,21,22^ It also allows for an intersectional assessment of indicators. For example, the child health-focused COVKID Scorecard found that though many states report on COVID-19 outcomes by age and race, a limited number of states report on the race of cases by age group (thereby allowing for an assessment of racial disparities among children).^8^ The sharing of case-delimited data allows users to pursue these more precise lines of inquiry.

A percent score was computed by dividing the sum of points awarded by the total number of indicators (N=135). Two sub-scores were also computed, a score for “overall” data available for each level of aggregation overall (5 data elements * 3 population aggregation units * 1 “overall” stratum = total score out of 15) and an equity indicator score for reporting across the remaining social strata (5 data elements * 3 population aggregation units * 8 social strata = total score out of 120).

We adjusted score denominators to take into account that reporting on some of the indicators, such as the cumulative total of deaths or hospitalizations, is less relevant in jurisdictions without any recorded cases or when case numbers are so low (i.e. n<5) that reporting may jeopardize patient confidentiality. When the total number of observations needed to estimate the indicator was less than 5, the indicators were removed from the denominator total. In that way, if one jurisdiction had not recorded any COVID-19 cases, for example, it was not penalized for not reporting on cases by age, sex, etc. Lastly, the following letter grades were associated with documented percent scores: 0% to 24% scores graded as “D”, 25% to 49% as “C”, 50% to 74% as “B”, and 75% to 100% as “A”.

## Results

Scores were estimated for each province and territory, and Canada overall (Figure 1, with detailed scores and sources in Supplementary File, Table 1). On average, approximately half (53%) of the data elements were available at the overall jurisdictional, health region, and local neighbourhood level, while approximately one in five (21%) equity-related data elements were available.

**Figure 1:**
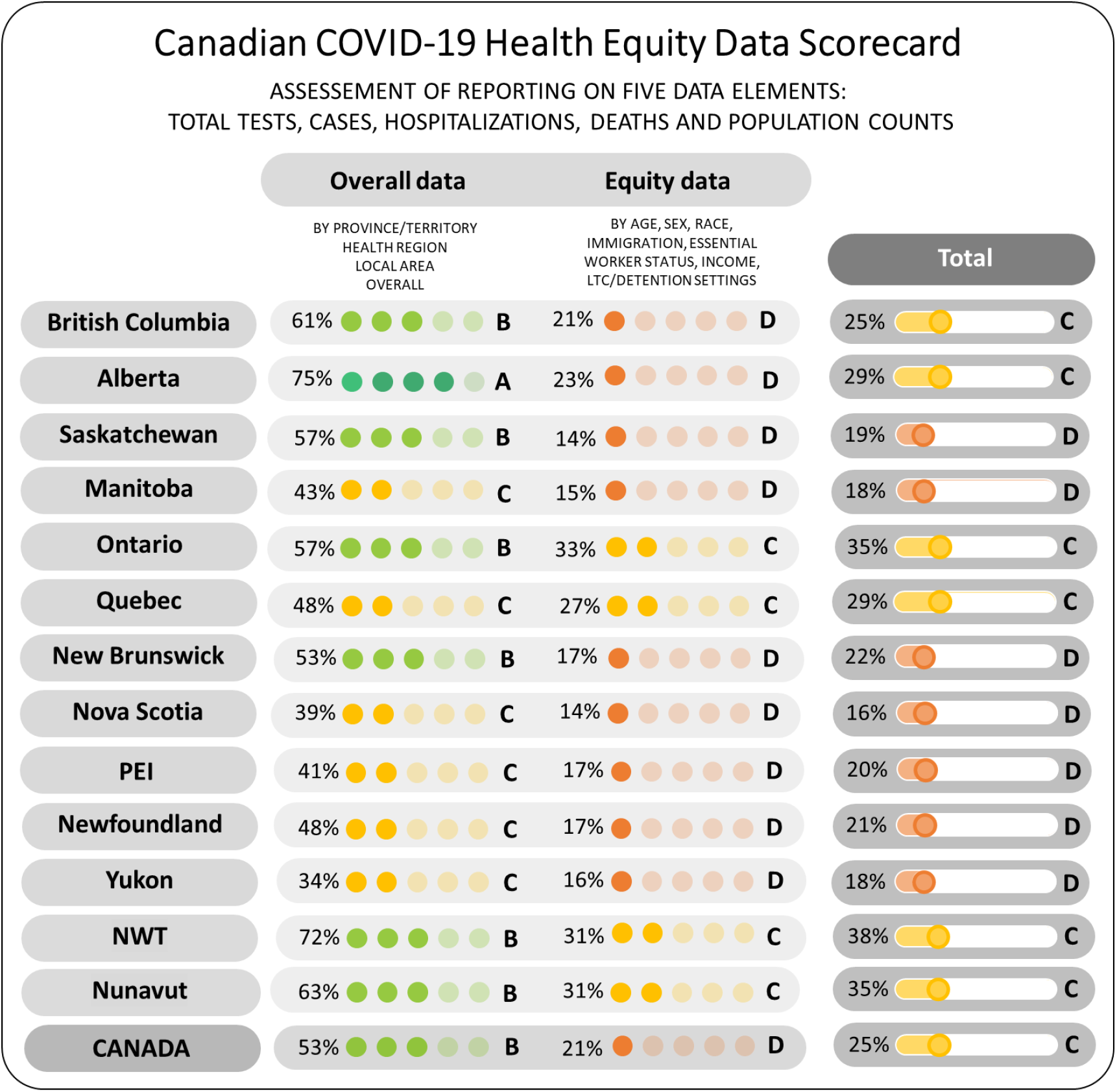
Total- and sub-scores (for reporting on overall and equity-related data) in Canada, by province and territory. Note: A grade of “D” was attributed to scores of 0% to 24%; “C” to scores of 25% to 49%; “B” to scores of 50% to 74%, and “A” to scores of 75% to 100%. “PEI” corresponds to Prince Edward Island, “NWT” to Northwest Territories.

### 3.1 Overall data reporting

#### 3.1.1 By provinces and territories

All provinces and territories reported on the total number of tests and cases (Figure 2a-b), with British Columbia and Alberta providing case-delimitated data for all cases observed (Figure 2b). Most jurisdictions also reported on the total number of hospitalizations that have occurred, however, these data were not provided by Nova Scotia, Newfoundland, and the Yukon (Figure 2c). All jurisdictions that had identified cases reported on the total number of deaths (Figure 2d). Alberta is the only province that reported on the outcomes (recovery or death) for each case, in a downloadable case-delimited format. Lastly, population denominators for all jurisdictions were available through the Canadian Census.

**Figure 2:**
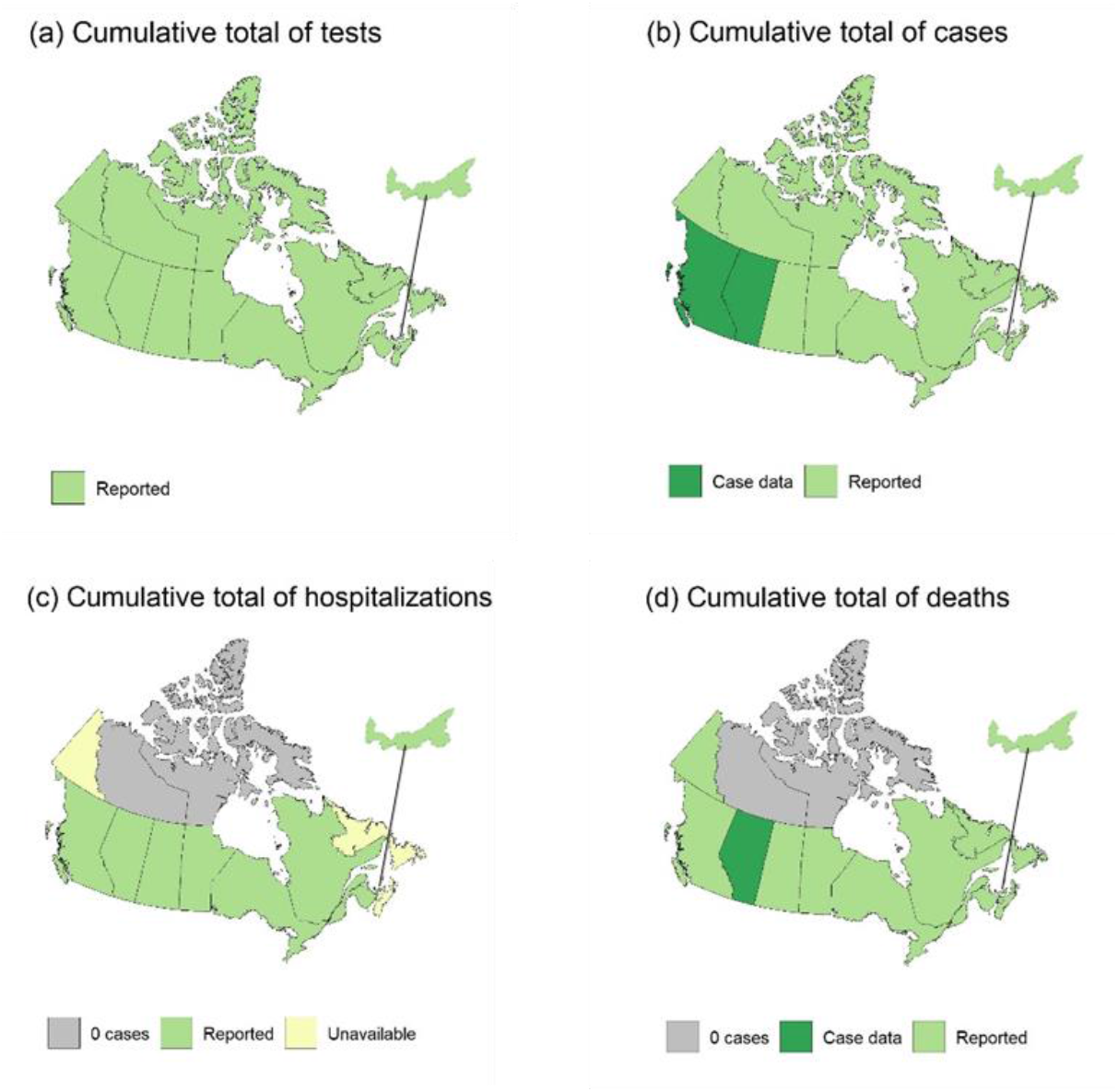
Overall, province- and territory-level reporting (data availability) on the cumulative total of tests, cases, hospitalizations, and deaths.

#### 3.1.2 By health region

The overall number of tests conducted per health region was only available for half of the jurisdictions (Figure 2a). Reporting on the total number of cases per health region was more complete, with British Columbia and Alberta both standing out as provinces that provide data on the region of residence for all identified cases (Figure 2b). Overall, most provinces that reported on overall hospitalizations (Figure 1c) also provided summaries of hospitalizations per health region (Figure 2c)—New Brunswick, Quebec, and Prince Edward Island were the exceptions to this rule. Except for Nova Scotia and the Yukon, data on deaths per health region were available for all jurisdictions reporting over five COVID-19 cases (Figure 2d). Lastly, population denominators for all health regions within jurisdictions were available from Statistics Canada’s Canadian Census.

#### 3.1.3 By Forward Sortation Area or local neighbourhood area equivalent

Though population denominators are made available by Statistics Canada for all Forward Sortation Areas in Canada, no province or territory reported on the overall number of tests or hospitalizations at this level of population aggregation. Alberta was the only jurisdiction to report on the number of cases and deaths (including the absence of the latter, n=0) for all local areas of residence—these were the only two data elements included in their reporting at the local area-level.

### 3.2 Equity-oriented reporting by social markers and vulnerable settings

#### 3.2.1 By age and sex

Information on population sizes by age and sex overall, and for regions and local areas is available through the Canadian Census. Of all the social markers studied, age and sex were the characteristics for which COVID-19 data reporting was most common. At the overall-level, almost all provinces reported on cases’ age and sex distribution—save for Newfoundland, which did not report sex-disaggregated case information (Figure 4). British Columbia and Alberta were the only two provinces that provided age and sex characteristics of all cases, in a case-delimited format. The Yukon, the only territory with over five recorded cases, did not report on cases’ age nor sex (Figure 4) at the territory or regional level.

**Figure 3:**
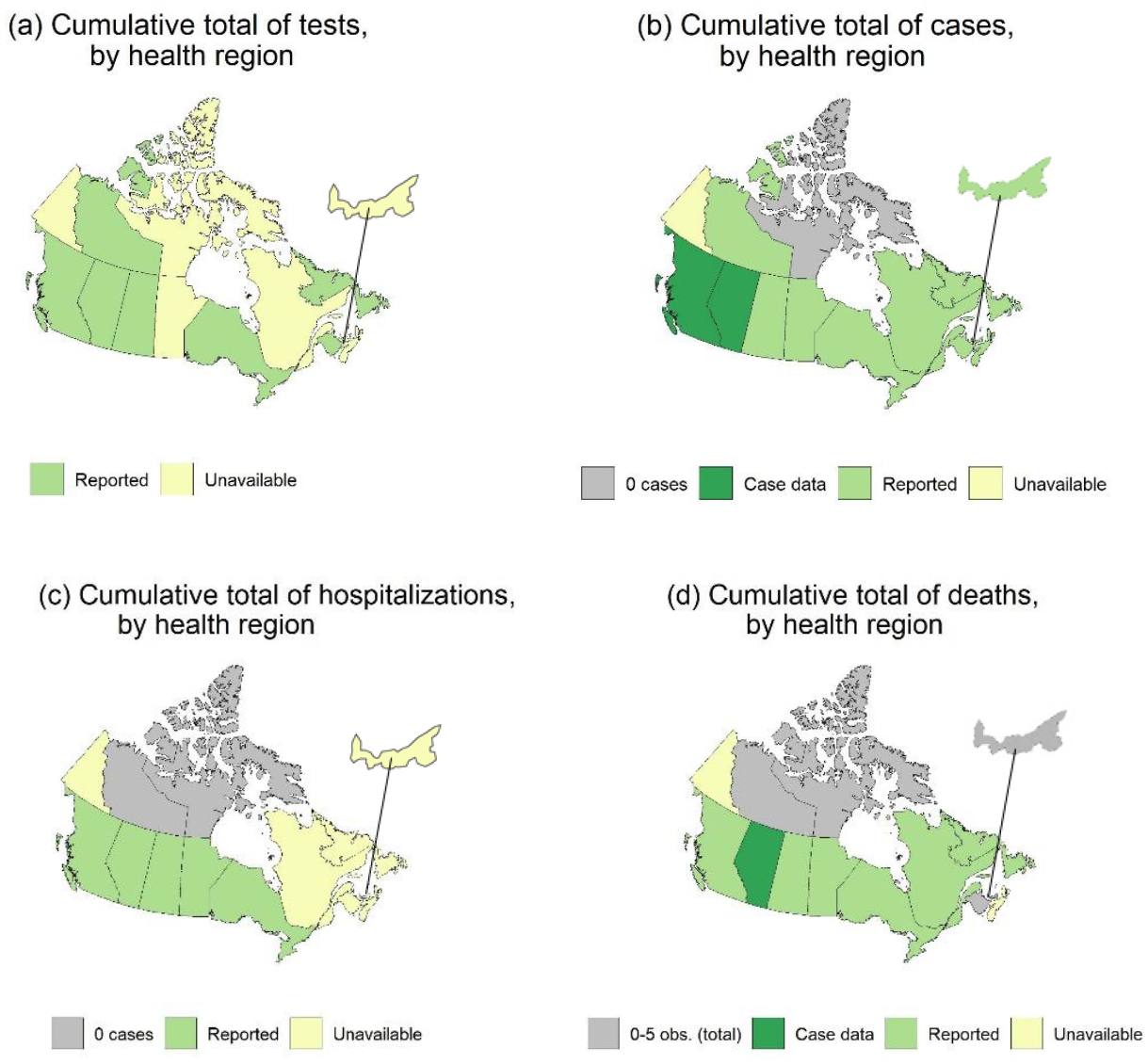
Reporting (data availability) on the cumulative total of tests, cases, hospitalizations, and deaths, overall, for each health region or health unit within the province/territory.

**Figure 4:**
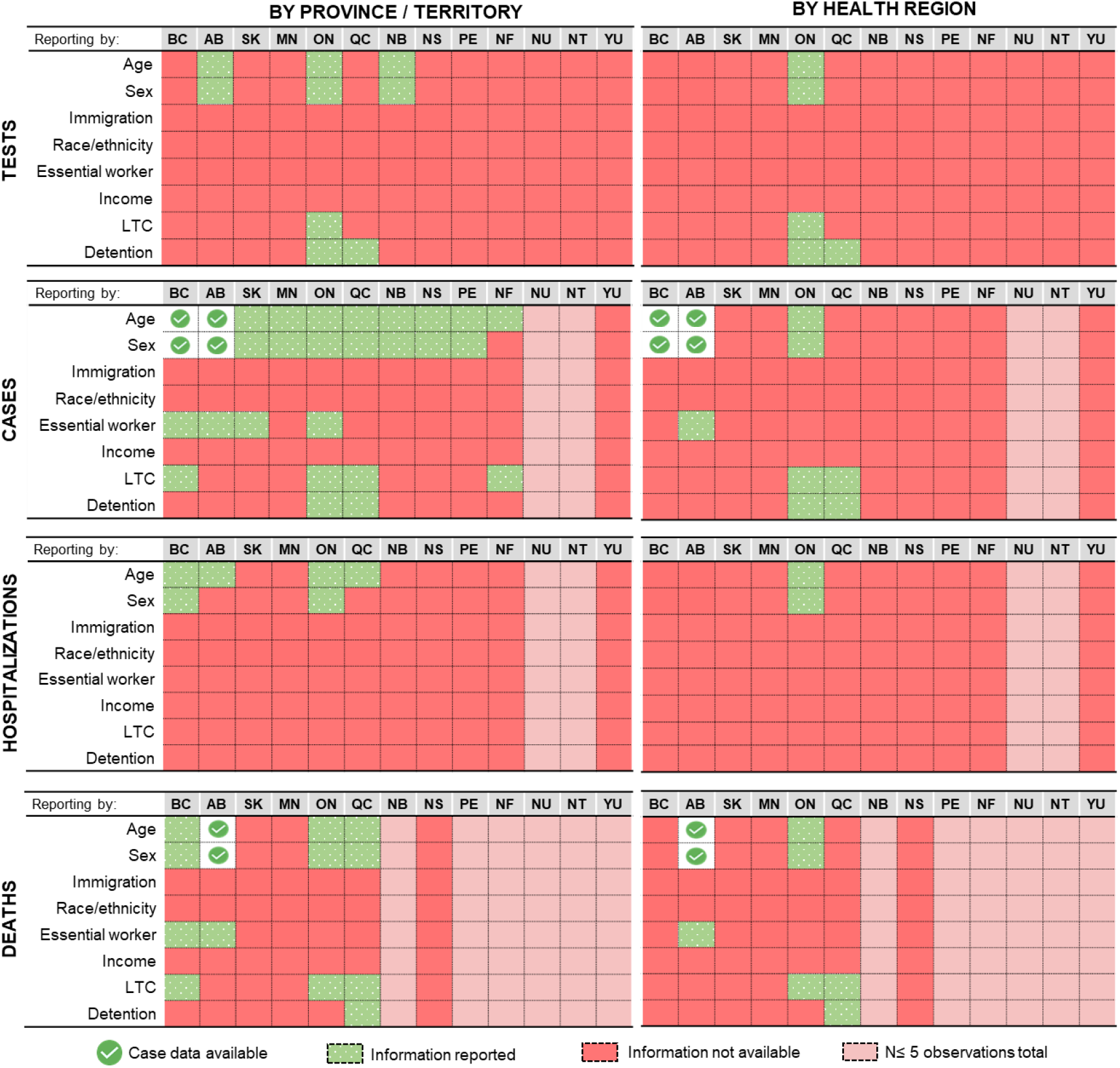
Reporting, by province/territory, and by health region, on the cumulative total of tests, cases, hospitalizations, and deaths by social markers and vulnerable settings. Acronyms: “BC” British Columbia, “AB” Alberta, “SK” Saskatchewan, “MN” Manitoba, “ON” Ontario, “QC” Quebec, “NB” New Brunswick, “NS” Nova Scotia, “PE” Prince Edward Island, “NF” Newfoundland, “NU” Nunavut, “NT” Northwest Territories, “YU” Yukon, “LTC” Long-term care settings.

In contrast, age- and sex-related information was sparser for testing, hospitalizations, and deaths across all jurisdictions overall and by health regions. Only Ontario consistently reported on all data elements by age and sex at the overall provincial and health region level (Figure 4).

#### 3.2.2 Immigration, race/ethnicity, income, essential worker status

Though information on population sizes by immigration status, race/ethnicity, and income are available for jurisdictions overall, and by region and local area are all available through the Canadian Census, no province or territory reported on any of the data elements according to these social markers (Figure 4).

At the overall level, on British Columbia, Alberta, Saskatchewan and Ontario provided information on cases among essential healthcare workers—with Alberta providing this information at both the province and regional levels (Figure 4). However, the total number of essential workers at the provincial or territorial level, or by region or local area were missing for all jurisdictions.

#### 3.2.3 Vulnerable settings (long-term care and detention facilities)

Ontario, Quebec, Newfoundland, British Columbia were the only jurisdictions that consistently provided detailed data on cases (and deaths) associated with long term care facilities—with Ontario and Quebec including listing precise facilities (that could be geolocated in local areas) that had or were experiencing outbreaks. In the latter provinces, however, the total number of tests and hospitalizations recorded for patients or staff in these settings was missing, and only Ontario provided data on the total number of beds per facility experiencing an outbreak—which could be used as a proxy for patient population size.

Quebec and Ontario were the only provinces that reported on the total number of cases for each provincial detention facility. Of these two, Quebec was the only province to report on the total number of tests, deaths, number of prisoners, and cases among staff per facility. Missing, however, was information on cases’ potential hospitalization status.

## Discussion

This paper provides the first summary of health equity-related COVID-19 data reporting in Canada. In Canada, information on cases and deaths was more complete than for tests, hospitalizations and population denominators for all indicators. Jurisdictions tended to report more completely on overall statistics than on information according to population sub-groups. The scan suggests that large gaps in reporting remain, even for more standard social disaggregation markers such as age and sex. Though relatively uncommon across the country, certain “best practices” in reporting emerged. For example, two provinces (Alberta and British Columbia) provided case-delimited data on cases for external users to study. Alberta was singular in reporting data elements for each of the three levels of geography: for the province overall, by health region, and by local area-level. Ontario and Quebec consistently provided detailed information for long term care settings, going as far as listing individual facilities that had or were experiencing outbreaks – which can enable the precise geo-location of facilities within neighbourhoods, for use in socio-spatial analyses of transmission risk. Lastly, though Ontario and Quebec both provided details on cases within provincial detention facilities, Quebec was alone in providing detailed information on COVID-19 tests, deaths, prisoner population size, and cases within staff populations per detention facilities. These examples set important precedents and guidance for other jurisdictions to follow, especially as emerging evidence suggests that if COVID-19 outcomes are properly examined across population subgroups, underlying inequities can be revealed and addressed.^3,4^

Heterogeneities in reporting observed across Canada are aligned with previous findings that public health surveillance infrastructures and capacities tend to vary across jurisdictions in Canada—which had been identified as an area of concern for pandemic planning and preparedness following the SARS outbreak in 2003.^23^ This variability in resources across jurisdictions may limit capacities to collect necessary social data and report on findings across settings or social markers. Exchange of promising practices—be it on equity-related reporting guidelines,^24^ questionnaires for social data collection,^17^ or data communication—may be beneficial to improve equity-related reporting in Canada. The scorecard approach presented here can be used for continued assessesments of COVID-19 surveillance reporting or adapted for use in future infectious disease outbreaks.

However, the scorecard approach used has certain limitations. For one, a restricted list of social marker indicators was used. Future expanded versions of an equity-oriented scorecard could assess COVID-19 outcome reporting according to indicators such as preferred language, year of immigration, disability status, sexual orientation, housing status, level of social support, gender, Indigenous identity, or education level.^17^ Second, by evaluating provincial or territorial reporting, this scorecard assessment did not address more detailed reporting efforts in smaller public health units. For instance, detailed neighbourhood-level reporting efforts have been made by Montreal Public Health^25^ and several public health units in Ontario.^26^ The present scan was restricted to provincial and territorial reporting in order to contrast between jurisdictions on a national scale. Future scans of best practices at the regional level may be warranted. Further, this scan excludes information sharing by federal bodies such as the Correctional Service of Canada’s reporting on cases within federal penitentiaries.^27^ Future reporting assessements asessing federal-level reporting may also be warranted.

## Conclusion

Though some “best practices” in health equity-oriented reporting were observed in Canada, equity data reporting is sparse and large gaps remain. Since jurisdictions that have explored potential social inequities in COVID-19 indicators have found stark gradients in outcomes across individual- and local-area level characteristics, the absence of reporting of data according to vulnerable settings or social markers may be concealing broader COVID-19-related inequities in Canada. The proposed scorecard format and examples of “best practices” identified herein can be used to guide surveillance and reporting during this pandemic and in and future ones.

## Data Availability

The data are provided as a supplementary file.

## Funding

AB receives postdoctoral funding from the Fonds de Recherche du Québec-Santé. AS is supported by the Canada Research Chair in Population Health Equity.

## Competing interests

None.

## Author contributions

AB, AP, and AS designed the scorecard framework. KW, HN, WB, and AB performed the environmental scan and collected the data for the study. AB drafted the manuscript, which was revised by AS, AP, KW, HN, and WB.

## Notes

### Competing Interest Statement

The authors have declared no competing interest.

### Funding Statement

AB receives postdoctoral funding from the Fonds de Recherche du Quebec-Sante. AS is supported by the Canada Research Chair in Population Health Equity.

### Author Declarations

Our paper uses publicly available, secondary, de-identified, ecological data, and as such is exempt from ethics review.

